# Persistent health inequalities over 20 years among adults with intellectual disabilities who display behaviours that challenge: Evidence from English primary care records

**DOI:** 10.64898/2025.12.17.25342459

**Authors:** Memta Jagtiani, Aws Sadik, Louise Marston, Shoumitro Deb, Dheeraj Rai, Bhathika Perera, Rohit Shankar, Juliette O’Connell, Angela Hassiotis

## Abstract

**Background:** Adults with intellectual disabilities who display behaviours that challenge (BtC) are more prone to poor health. This study seeks to evidence the long-term health outcomes for this population.

**Methods:** We conducted a longitudinal cohort study of adults with intellectual disabilities aged 18+ years in England using data from Clinical Practice Research Datalink Aurum (01/2003–12/2023) linked to Hospital Episode Statistics and Office for National Statistics. Main outcome measures were Annual Health Checks (AHCs), GP referrals, emergency visits, outpatient attendance, inpatient admissions, and mortality.

**Results:** Among 83,166 adults with intellectual disabilities (mean age 38.6 years), 18.5% had a record of BtC with similar sociodemographic distributions to those without BtC but higher rates of physical and mental health comorbidities and uptake of AHCs. 72.5% of participants with BtC were receiving psychotropic medication(s). Adults with BtC had higher rates of mental health outpatient attendance (OR: 1.42, 95% CI: 1.33 to 1.52) and inpatient admissions (IRR: 1.19, 95% CI: 1.09 to 1.29) but consistently lower rates of physical health outpatient attendance (IRR: 0.81, 95% CI: 0.78 to 0.84) and inpatient admissions (IRR: 0.77, 95% CI: 0.74 to 0.79), after adjusting for demographic and clinical characteristics. BtC was not associated with mortality after adjustment for comorbidities (HR: 0.97, 95% CI: 0.93 to 1.00).

**Conclusion:** This longitudinal study not only corroborated the markedly elevated burden of physical and mental health comorbidities among individuals displaying BtC but also indicated that repeated efforts to improve health outcomes have yielded minimal measurable benefit over time. The apparent absence of progress is likely underpinned by a combination of insufficiently effective or poorly tailored interventions and wider systemic constraints that limit the capacity of services to respond to the complex needs of this population.

## INTRODUCTION

Intellectual disabilities is a neurodevelopmental disorder characterised by significant limitations in intellectual functioning and adaptive behaviour with onset during the developmental period [1]. Individuals with intellectual disabilities often display behaviours that challenge (BtC), including verbal or physical aggression, self-injury and destruction to property. These behaviours are associated with poor quality of life, social exclusion, and increased risk of psychiatric hospitalisation [2].

Multiple interventions including psychotropic medications and psychosocial interventions have been trialled in the management and prevention of BtC. Groves et al (2023) in a meta-analysis of 82 reports of clinical trials found no superiority of pharmacological vs non-pharmacological interventions for BtC and its specific typologies [3]. Psychotropic medications, especially antipsychotics, are frequently prescribed off-licence to manage BtC particularly where there may be risk to self or others [4, 5]. In England, national guidelines recommend that antipsychotics be used only in the short term when risks are high, and alongside non-pharmacological interventions [6]. However, in practice, adults with intellectual disabilities who display BtC are more likely to be prescribed antipsychotics long term [7], the drivers of which are underpinned by multiple factors including clinical presentation, efficacy of service approaches, or even caregiver preferences [8, 9]. Since 2008, national initiatives such as annual health checks (AHCs) for people with intellectual disabilities aged 14 years and over have aimed to improve the mental and physical health of this population and reduce health inequalities [10]. Buszewicz et al. (2014) found that AHCs were associated with improved detection of health conditions, increased delivery of health promotion, and reduced preventable hospital admissions [11].

### Gaps in literature

Longitudinal associations between BtC, therapeutic input and health outcomes among adults with intellectual disabilities remain under-researched [12]. Given substantial uncertainties of the long-term outcomes of the relapsing-remitting nature of BtC, sources such as large routinely collected data may offer a unique opportunity to assess outcomes over time, generate system-level insights into patterns of care, and monitor prescribing practices and potential inequalities within this vulnerable group.

### Aim

To investigate and compare clinical outcomes across primary and secondary healthcare use in adults with intellectual disabilities with and without BtC aged 18 years and over.

### Objectives

1. To describe the cohort of adults with intellectual disabilities in terms of sociodemographic and clinical characteristics.
2. To estimate the prevalence of psychotropic prescribing and non-pharmacological interventions in the identified cohort.
3. To estimate and compare patterns of AHCs and GP referrals for further assessment and/or treatment.
4. To determine the proportion of individuals with and without BtC identified in the cohort who subsequently had any hospital contact, i.e., attendance, admissions and mortality.
5. To examine whether BtC is associated with mortality and any hospital contact, i.e., attendance and admissions, while adjusting for age, ethnic group, deprivation index, and diagnostic comorbidities.

## METHODS

### Participants

We identified a longitudinal cohort of patients with intellectual disabilities who were registered with a primary care practice in England between 2003 to 2023, from the Clinical Practice Research Datalink (CPRD) Aurum. CPRD Aurum is representative of the population [13] and contains electronic medical records for over 16 million patients [14].

A set of clinical codes related to intellectual disabilities, health comorbidities and neurodevelopmental or psychiatric conditions were collated from CPRD Aurum’s code browser between June and August 2024. These were used to extract the records of adults with intellectual disabilities (code lists available on request).

The eligibility criteria were having a record of intellectual disabilities and linked data (see below), registered with an up-to-standard general practice and aged 18 years or older at or after 1^st^ January 2003 until 31^st^ December 2022 to allow for at least a year of follow-up. Individual patient records were analysed from cohort inception or registration start date or year they turned 18 years old (whichever was the latest) until the end of data study: 31^st^ December 2023 or registration end date or patient death date or the year they turned 100 years old (whichever was the earliest).

### Data Linkage

In addition to CPRD Aurum, this study used linked data from Office for National Statistics (ONS) death registration data, Hospital Episode Statistics (HES) secondary care datasets, including HES Admitted Patient Care (HES APC), HES Outpatient (HES OP) data and HES Accident and Emergency (HES A&E) data. All the linked datasets covered the years of the cohort (2003 to 2023) except HES A&E data which only covered the period 2007 to 2020. It was replaced by the Emergency Care Data Set (ECDS) from April 2020 but is not yet linked to CPRD. HES data is not available for everyone as HES does not link to all practices, therefore the cohort size is smaller after data linkage. Linkage of CPRD to HES datasets is carried out by a trusted third-party (NHS Digital) to maintain patient confidentiality.

### Ethical approval

The authors assert that all procedures contributing to this work comply with the ethical standards of the relevant national and institutional committees on human experimentation and with the [Helsinki Declaration](https://www.wma.net/policies-post/wma-declaration-of-helsinki-ethical-principles-for-medical-research-involving-human-subjects/) of 1975, as revised in 2013. All procedures involving human subjects/patients were approved by the Research Ethics Committee of the Health Research Authority to support research using anonymised patient data (reference: 21/EM/0265).

### Consent

The CPRD electronic Research Application Portal (eRAP) was used to request access to the CPRD data and obtain approval for the study (protocol reference ID: 23_002605). Patients can opt out of having their de-identified data shared via CPRD. Similarly, all linked datasets used the opt-out consent model.

### Exposure

The exposure for this study was adults with intellectual disabilities who display BtC. Related SNOMED CT codes were identified from CPRD Aurum’s medical code dictionary. Code lists were also obtained and adapted from other researchers and institutions (e.g., LSHTM Data Compass) and verified by clinicians in the research team.

### Outcomes

We were interested in:

1. Annual health checks
2. GP referrals
3. Emergency visits (HES A&E)
4. Physical and mental health outpatient attendance (HES OP)
5. Physical and mental health inpatient admissions (HES APC)
6. All-cause mortality (CPRD-ONS)

The segregation of physical and mental health inpatient admissions and outpatient attendance was achieved by grouping the 10^th^ revision of the International Classification of Diseases (ICD-10) codes, where codes that started with F indicated mental ill-health and all other codes were classified as physical ill-health. GP referrals were categorised as physical health referral, mental health referral or unspecified referral. The most used codes for the latter are “referred to hospital” or “hospital referral”.

### Potential confounders

We included sex, ethnicity and Index of Multiple Deprivation (IMD) quintile as potential sociodemographic confounders. Sex and ethnicity were reported as recorded in the patient’s Electronic Health Record (EHR). Ethnicity, recorded in both CPRD and HES, was reported as defined in either source of data at any time point [15] and grouped according to the UK 2011 Census Ethnic Group categories [16]: ‘Asian’, ‘Black’, ‘Mixed’, ‘White’ or ‘Other’. Where conflict existed between CPRD and HES, we prioritised the CPRD classification because HES ethnicity recording is less accurate [17]. IMD was based on patient postcode and, where this was missing, the primary care practice postcode was used to indicate IMD quintile.

We investigated mental and physical comorbidities as potential confounders as they are key drivers of mortality and hospitalisations. Code lists for mental and physical comorbidities were defined based on clinical input and other sources (https://github.com/Exeter-Diabetes/CPRD-Codelists).

### Statistical analysis

We conducted descriptive analyses to characterise the study cohort in terms of demographic variables, IMD, physical and mental health comorbidities, and the types of interventions received.

The cohort was stratified by the presence or absence of BtC. Categorical variables were summarised using frequencies and percentages, while continuous variables such as age and follow-up time were described using means and standard deviations. Follow-up time was calculated from the date of cohort entry to when the participant left the cohort. Patient-years were computed to reflect the total time at risk contributed by individuals over the follow-up period. The proportion of patients who received no intervention, non-pharmacological interventions only (e.g., behavioural or psychological interventions – code lists available on request), psychotropic medications only (antidepressants, antipsychotics, anxiolytics, sedatives and hypnotics, antiparkinsonian agents, mood stabilisers, ADHD drugs, and antidementia drugs), and both types of interventions at any time were also computed.

For all outcomes we included the same variables in the same order adjusting for demographic (age, sex, ethnicity, IMD quintile) and clinical status (neurodevelopmental conditions, mental and physical health comorbidities). Some data were missing for ethnicity and IMD. All analyses used complete case analysis and were conducted using R version 4.2.5.

#### Mortality

We used Cox proportional hazards regression to assess the relationship between BtC and all-cause mortality. Time-to-event was calculated from age 18 for all to the earliest date of death or where participants were censored. We reported hazard ratios and corresponding 95% confidence intervals. Kaplan-Meier survival curves were plotted to visualise unadjusted survival between BtC and non-BtC groups.

#### Service Use

For physical health outpatient attendance and physical and mental health hospital admissions, we applied negative binomial regression models because of overdispersion in the outcomes, with an offset of 1/time in the cohort to account for the different lengths of time participants were in the cohort. We reported the incidence rate ratios and corresponding 95% confidence intervals.

We used logistic regression to model the odds of having at least one mental health outpatient attendance as a binary outcome because there were few people with mental health outpatient attendance recorded and the negative binomial models did not converge. We reported the odds ratios and corresponding 95% confidence intervals.

## RESULTS

### Description of the cohort

Figure 1 illustrates how the final analytical sample (n = 83,166) was derived.

**Figure 1:**
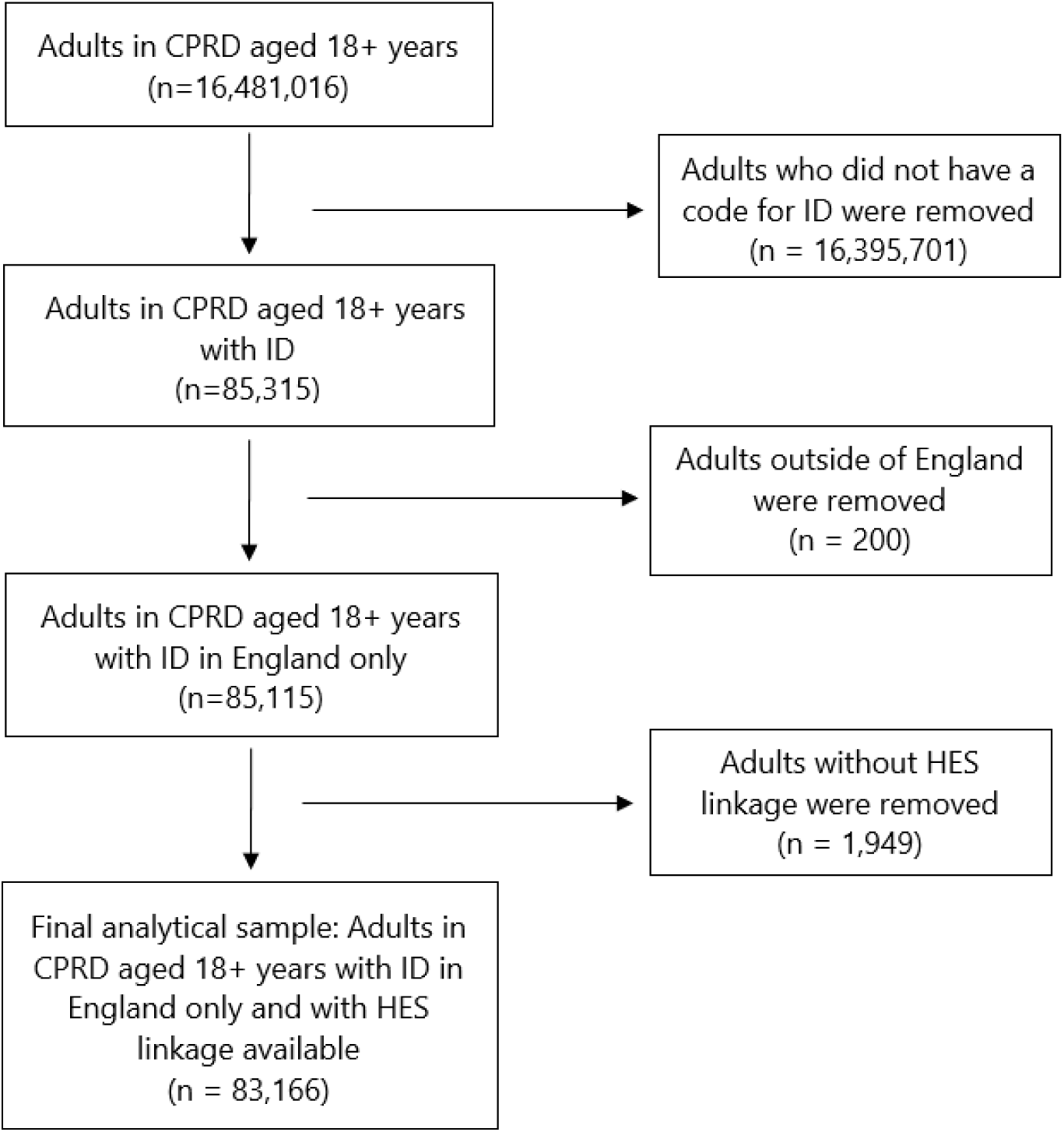
Flow diagram of participants

The total number of patient-years was 540,509 with a mean follow-up of 6.5 years (SD=5.0). The mean age at cohort entry was 38.6 years. Out of the total cohort (n=83,166), 18.5% (n=15,368) had a record of BtC. Males made up 57.3% of the cohort, with similar sex distributions across BtC and non-BtC groups. Most of the cohort was White (82.6%). The distribution across IMD quintiles was comparable between groups, with 23.9% (n=19,845) residing in the most deprived areas (Table 1).

**Table 1:** Sociodemographic and clinical profile of the cohort with intellectual disabilities from 2003 to 2023

|  | **Overall, n (%)** | **BtC, n (%)** | **No BtC, n (%)** |
| --- | --- | --- | --- |
| **Cohort, n (row %)** | 83,166 (100.0) | 15,368 (18.5) | 67,798 (81.5) |
| **Sex, n (column %):** |  |  |  |
| Females | 35,480 (42.7) | 6,365 (41.4) | 29,115 (42.9) |
| Males | 47,686 (57.3) | 9,003 (58.6) | 38,683 (57.1) |
| **All-cause mortality** |  |  |  |
| Died between 01/01/2004 and 31/12/2023 | 22,933 (27.6) | 5,144 (33.5) | 17,789 (26.2) |
| Age at death, mean (SD) for those who died | 62.8 (16.0) | 64.1 (14.8) | 62.5 (16.3) |
| Age at death, median for those who died | 64 | 65 | 64 |
| **Age:** |  |  |  |
| Age at entry, mean (SD) | 38.6 (17.8) | 41.2 (17.7) | 38.1 (17.8) |
| **Ethnicity:** |  |  |  |
| White | 68,693 (82.6) | 13,247 (86.2) | 55,446 (81.8) |
| Black | 2,957 (3.6) | 549 (3.6) | 2,408 (3.6) |
| Asian | 3,158 (3.8) | 594 (3.9) | 2,564 (3.8) |
| Mixed and Other | 1,288 (1.5) | 190 (1.2) | 1,098 (1.6) |
| Missing | 7,070 (8.5) | 788 (5.1) | 6,282 (9.3) |
| **Index of Multiple Deprivation:** |  |  |  |
| 1 – least deprived | 8,736 (10.5) | 1,534 (10.0) | 7,202 (10.6) |
| 2 | 11,675 (14.0) | 2,293 (14.9) | 9,382 (13.8) |
| 3 | 14,221 (17.1) | 2,811 (18.3) | 11,410 (16.8) |
| 4 | 16,026 (19.3) | 2,880 (18.7) | 13,146 (19.4) |
| 5 – most deprived | 19,845 (23.9) | 3,479 (22.6) | 16,366 (24.1) |
| Missing | 12,663 (15.2) | 2,371 (15.4) | 10,292 (15.2) |
| **Physical health comorbidities:** |  |  |  |
| High cholesterol | 24,403 (29.3) | 6,070 (39.5) | 18,333 (27.0) |
| Constipation | 16,084 (19.3) | 5,331 (34.7) | 10,753 (15.9) |
| Type 2 Diabetes | 8,070 (9.7) | 1,830 (11.9) | 6,240 (9.2) |
| Epilepsy | 17,281 (20.8) | 4,707 (30.6) | 12,574 (18.5) |
| High blood pressure | 20,290 (24.4) | 4,878 (31.7) | 15,412 (22.7) |
| Overweight and obese | 27,739 (33.4) | 6,920 (45.0) | 20,819 (30.7) |
| Severe mobility difficulties | 9,388 (11.3) | 3,309 (21.5) | 6,079 (9.0) |
| **Mental health comorbidities:** |  |  |  |
| Anxiety | 9,434 (11.3) | 3,097 (20.2) | 6,337 (9.3) |
| Lifetime record of dementia | 4,250 (5.1) | 1,331 (8.7) | 2,919 (4.3) |
| Depression | 14,458 (17.4) | 5,605 (36.5) | 8,853 (13.1) |
| **Severe mental illness:** |  |  |  |
| Lifetime record of bipolar disorder | 16,927 (20.4) | 6,411 (41.7) | 10,516 (15.5) |
| Lifetime record of psychoses and schizophrenia | 7,413 (8.9) | 2,753 (17.9) | 4,660 (6.9) |
| **Neurodevelopmental disorders:** |  |  |  |
| Lifetime record of ADHD | 4,402 (5.3) | 1,056 (6.9) | 3,346 (4.9) |
| Lifetime record of Autism | 12,221 (14.7) | 3,123 (20.3) | 9,098 (13.4) |
| **Interventions:** |  |  |  |
| None | 33,567 (40.4) | 2,486 (16.2) | 31,081 (45.8) |
| At least 1 non-pharmacological intervention only | 1,301 (1.6) | 266 (1.7) | 1,035 (1.5) |
| At least 1 psychotropic medication only | 44,741 (53.8) | 11,144 (72.5) | 33,597 (49.6) |
| Both | 3,557 (4.3) | 1,472 (9.6) | 2,086 (3.1) |

The BtC group had higher rates of physical comorbidities than those in the non-BtC group across all conditions examined. A similar pattern was observed for mental health conditions, with notably higher prevalence of depression (36.5% vs. 13.1%), anxiety (20.2% vs. 9.3%), bipolar disorder (41.7% vs. 15.5%), psychosis or schizophrenia (17.9% vs. 6.9%), and autism (20.3% vs. 13.4%) in the BtC group.

Mortality was higher among those with BtC (33.5% vs. 26.2%). Mean age at death was slightly higher in the BtC group compared to the non-BtC group (64.1 vs. 62.5).

More than half of the cohort (53.8%) were recorded as receiving at least one psychotropic medication only, with this being especially common in the BtC group, where 72.5% were prescribed any psychotropic medication. Notably, 40.4% of the cohort did not receive psychotropic medication(s) or non-pharmacological intervention(s). In the BtC group, 9.6% were recorded as receiving both psychotropic medication(s) and non-pharmacological intervention(s), compared to 3.1% in the non-BtC group.

Stratified by intervention type (Table 2), those in the BtC group have higher mortality if receiving both interventions (37.0%) or at least one psychotropic medication (34.8%), compared to those receiving no intervention (26.3%) or at least one non-pharmacological intervention (25.1%). Between males and females, the largest difference was observed among those who had a record of at least one non-pharmacological intervention (65% males vs. 35% females). There was little difference between ethnicities in terms of interventions received.

**Table 2:**
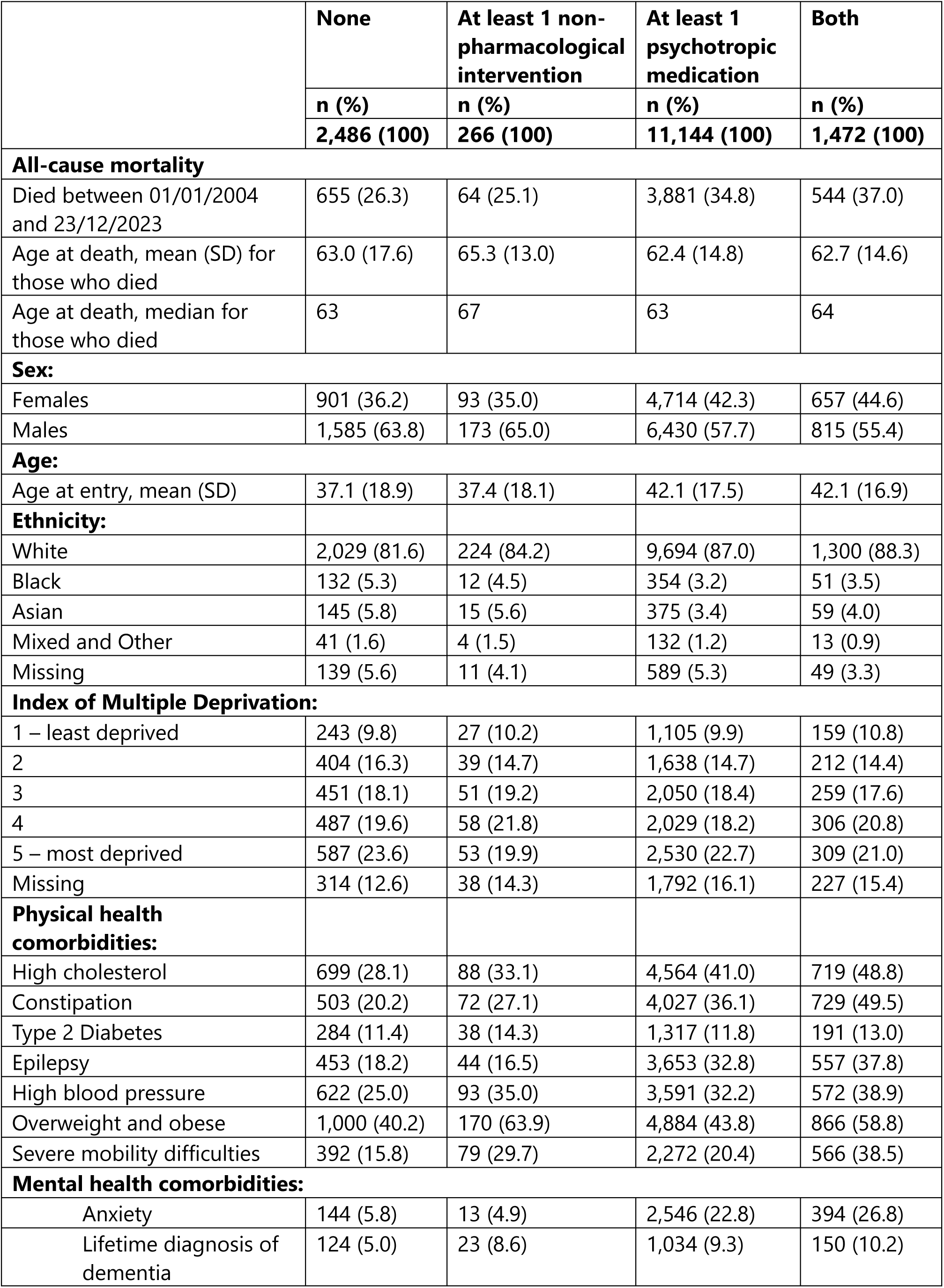
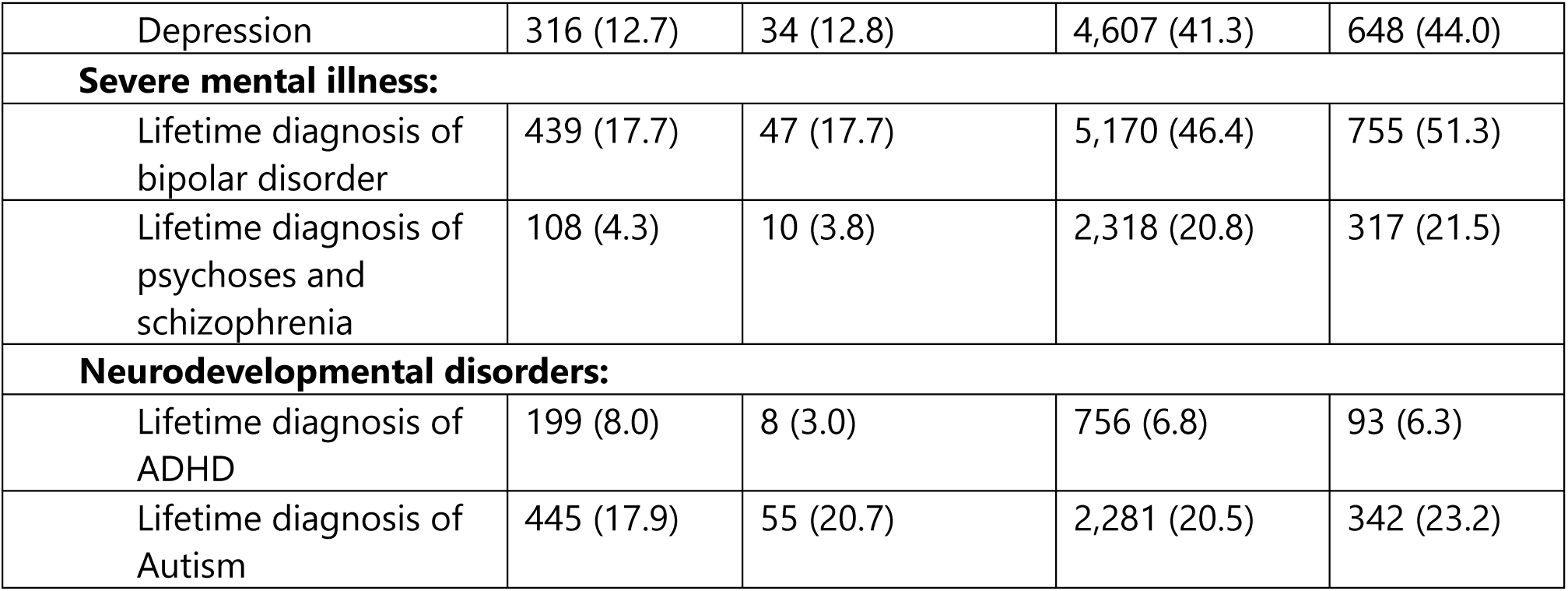
Sociodemographic and clinical profile of the BtC cohort (n = 15,368) by intervention type

|  | None | At least 1 non-pharmacological intervention | At least 1 psychotropic medication | Both |
| --- | --- | --- | --- | --- |
|  | n (%) | n (%) | n (%) | n (%) |
|  | 2,486 (100) | 266 (100) | 11,144 (100) | 1,472 (100) |
| **All-cause mortality** |  |  |  |  |
| Died between 01/01/2004 and 23/12/2023 | 655 (26.3) | 64 (25.1) | 3,881 (34.8) | 544 (37.0) |
| Age at death, mean (SD) for those who died | 63.0 (17.6) | 65.3 (13.0) | 62.4 (14.8) | 62.7 (14.6) |
| Age at death, median for those who died | 63 | 67 | 63 | 64 |
| **Sex:** |  |  |  |  |
| Females | 901 (36.2) | 93 (35.0) | 4,714 (42.3) | 657 (44.6) |
| Males | 1,585 (63.8) | 173 (65.0) | 6,430 (57.7) | 815 (55.4) |
| **Age:** |  |  |  |  |
| Age at entry, mean (SD) | 37.1 (18.9) | 37.4 (18.1) | 42.1 (17.5) | 42.1 (16.9) |
| **Ethnicity:** |  |  |  |  |
| White | 2,029 (81.6) | 224 (84.2) | 9,694 (87.0) | 1,300 (88.3) |
| Black | 132 (5.3) | 12 (4.5) | 354 (3.2) | 51 (3.5) |
| Asian | 145 (5.8) | 15 (5.6) | 375 (3.4) | 59 (4.0) |
| Mixed and Other | 41 (1.6) | 4 (1.5) | 132 (1.2) | 13 (0.9) |
| Missing | 139 (5.6) | 11 (4.1) | 589 (5.3) | 49 (3.3) |
| **Index of Multiple Deprivation:** |  |  |  |  |
| 1 – least deprived | 243 (9.8) | 27 (10.2) | 1,105 (9.9) | 159 (10.8) |
| 2 | 404 (16.3) | 39 (14.7) | 1,638 (14.7) | 212 (14.4) |
| 3 | 451 (18.1) | 51 (19.2) | 2,050 (18.4) | 259 (17.6) |
| 4 | 487 (19.6) | 58 (21.8) | 2,029 (18.2) | 306 (20.8) |
| 5 – most deprived | 587 (23.6) | 53 (19.9) | 2,530 (22.7) | 309 (21.0) |
| Missing | 314 (12.6) | 38 (14.3) | 1,792 (16.1) | 227 (15.4) |
| **Physical health comorbidities:** |  |  |  |  |
| High cholesterol | 699 (28.1) | 88 (33.1) | 4,564 (41.0) | 719 (48.8) |
| Constipation | 503 (20.2) | 72 (27.1) | 4,027 (36.1) | 729 (49.5) |
| Type 2 Diabetes | 284 (11.4) | 38 (14.3) | 1,317 (11.8) | 191 (13.0) |
| Epilepsy | 453 (18.2) | 44 (16.5) | 3,653 (32.8) | 557 (37.8) |
| High blood pressure | 622 (25.0) | 93 (35.0) | 3,591 (32.2) | 572 (38.9) |
| Overweight and obese | 1,000 (40.2) | 170 (63.9) | 4,884 (43.8) | 866 (58.8) |
| Severe mobility difficulties | 392 (15.8) | 79 (29.7) | 2,272 (20.4) | 566 (38.5) |
| **Mental health comorbidities:** |  |  |  |  |
| Anxiety | 144 (5.8) | 13 (4.9) | 2,546 (22.8) | 394 (26.8) |
| Lifetime diagnosis of dementia | 124 (5.0) | 23 (8.6) | 1,034 (9.3) | 150 (10.2) |
| Depression | 316 (12.7) | 34 (12.8) | 4,607 (41.3) | 648 (44.0) |
| **Severe mental illness:** |  |  |  |  |
| Lifetime diagnosis of bipolar disorder | 439 (17.7) | 47 (17.7) | 5,170 (46.4) | 755 (51.3) |
| Lifetime diagnosis of psychoses and schizophrenia | 108 (4.3) | 10 (3.8) | 2,318 (20.8) | 317 (21.5) |
| **Neurodevelopmental disorders:** |  |  |  |  |
| Lifetime diagnosis of ADHD | 199 (8.0) | 8 (3.0) | 756 (6.8) | 93 (6.3) |
| Lifetime diagnosis of Autism | 445 (17.9) | 55 (20.7) | 2,281 (20.5) | 342 (23.2) |

Individuals who were overweight and obese also had consistently higher proportions of all three combinations of interventions than those with other physical comorbidities. High cholesterol (48.8%), constipation (49.5%), and high blood pressure (38.9%) were especially common in the group that received both interventions, as were all mental health comorbidities, notably, severe mental illnesses (i.e., bipolar disorder (51.3%); psychosis/schizophrenia (21.5%). The small proportion of recorded non-pharmacological interventions is likely to be attributable to underreporting.

### Association between severe mental illness, BtC and prescribing of antipsychotics

Among the 24,699 adults with intellectual disabilities who were prescribed antipsychotics, 13,405 (54.3%) did not have a diagnosis of severe mental illness. Of the 15,368 individuals with a record of BtC, 8,096 (52.7%) received antipsychotic medication, of whom 4,698 (58.0%) had a diagnosis of severe mental illness. Finally, among those who were prescribed antipsychotics, 10,007 (40.5%) had no record of severe mental illness or BtC (Figure 2).

**Figure 2:**
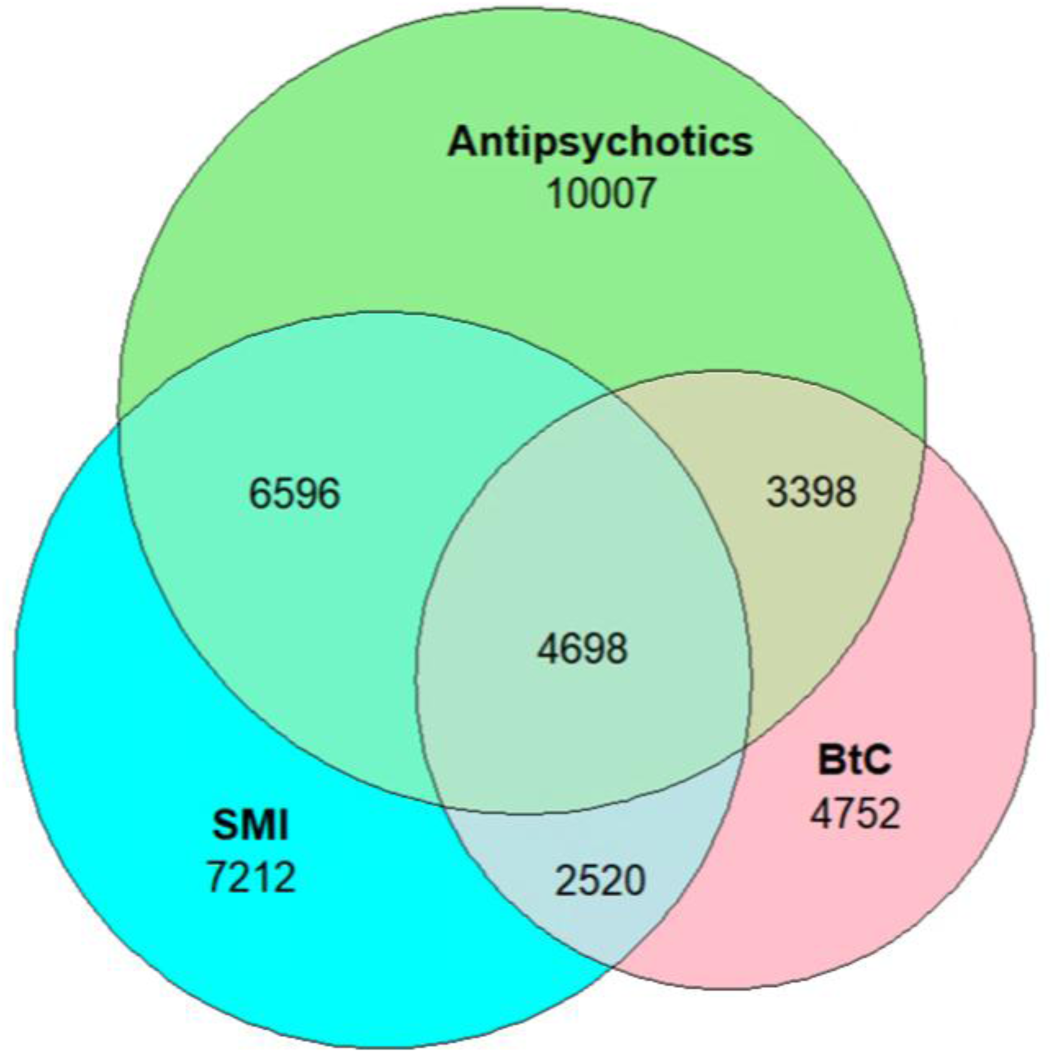
Associations between lifetime record of severe mental illness, BtC and prescription of antipsychotics Outcomes:

### Outcomes

#### 1. Annual Health Checks

Figure 3 shows the prevalence of AHCs in the cohort by various denominator groups: overall (i.e., everyone in the cohort), everyone except patients who had at least one exception code and no AHCs, patients on the intellectual disability register at any time and by BtC status. Exception reporting allows GP practices to exclude patients from specific indicators or clinical domains for reasons beyond their control [18].

**Figure 3:**
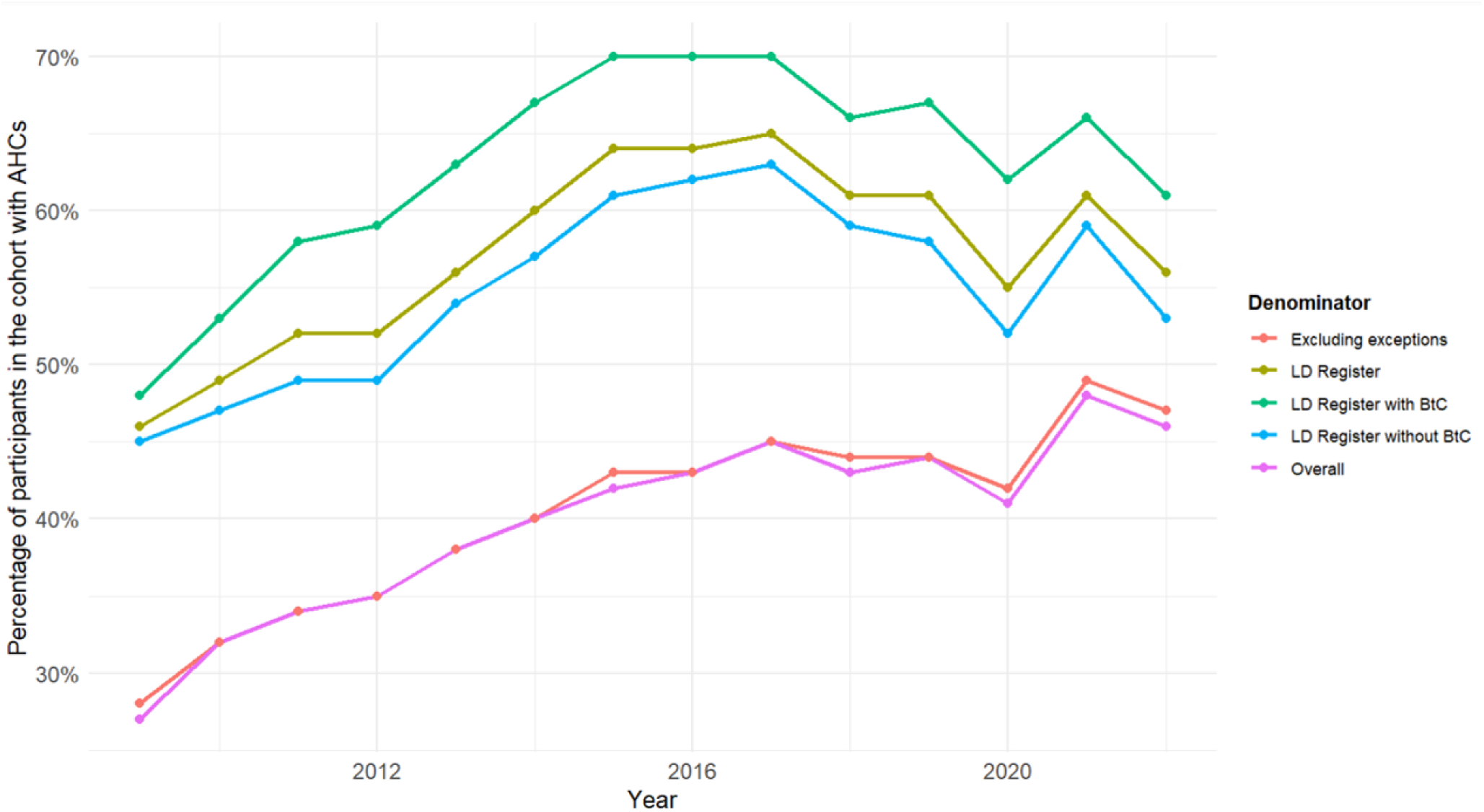
Time trends of annual health checks by BtC and register* status *It signifies the mandated recording of people with intellectual disabilities who attend primary care in England to support better healthcare access.

The percentage of individuals receiving AHCs increased from 2009 (27.4%) to 2017 (44.6%), followed by a gradual decline. A notable drop in AHCs occurred in 2020, likely due to the COVID-19 pandemic, but recovered after that. Trends were similar among the group with at least one exception code and no AHC.

AHC rates were consistently higher among patients who displayed BtC and were in the intellectual disabilities register. Uptake peaked at 69.8% in 2017 and despite COVID-19 disruptions, it remained above 60% through 2022. While AHC rates for those without BtC increased between 2009 to 2017, they were lower than the BtC group.

#### 2. GP referrals for further assessments and/or intervention

GP referrals covered data from 2009 onwards. The highest proportion of GP referrals were for physical health compared to mental health and unspecified reasons. Mental health referrals were the lowest among the three types, with a range of 3.6%-6.5% during the cohort period (Figure 4).

**Figure 4:**
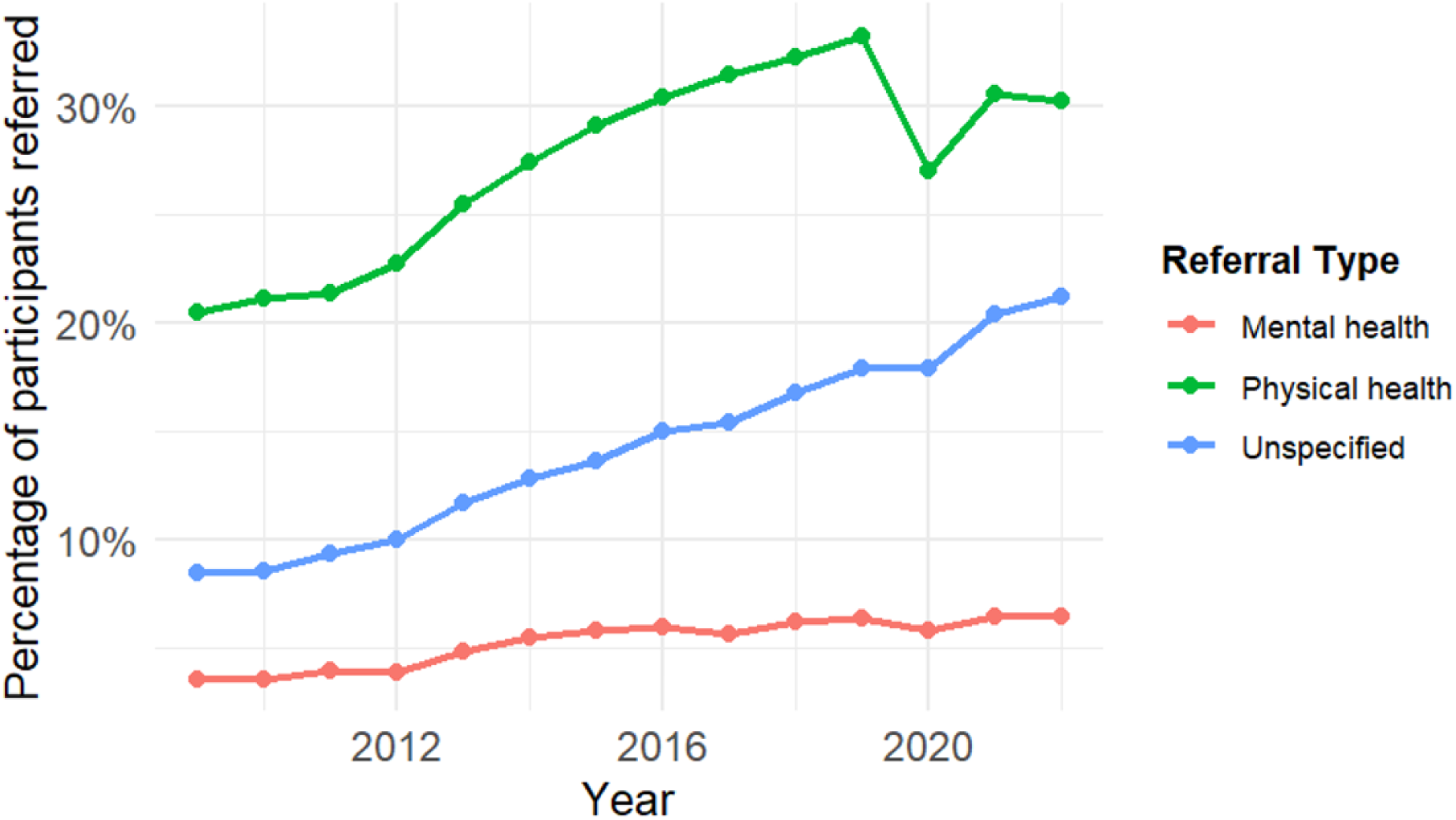
Time trends of GP referrals in the cohort

#### 3. All-cause Accident and Emergency attendance

Figure 5 shows the percentage of patients with at least one Accident and Emergency (A&E) attendance per year, stratified by BtC status. Patients with BtC had consistently lower A&E usage compared to those without BtC. The highest attendance rate for patients with BtC was 68% in 2019, followed by a drop to 30% in 2020 compared with rates of attendance in patients without BtC (83% in 2019, then dropping to 36% in 2020). These rates could reflect the crossover to the new system of recording A&E attendances.

**Figure 5:**
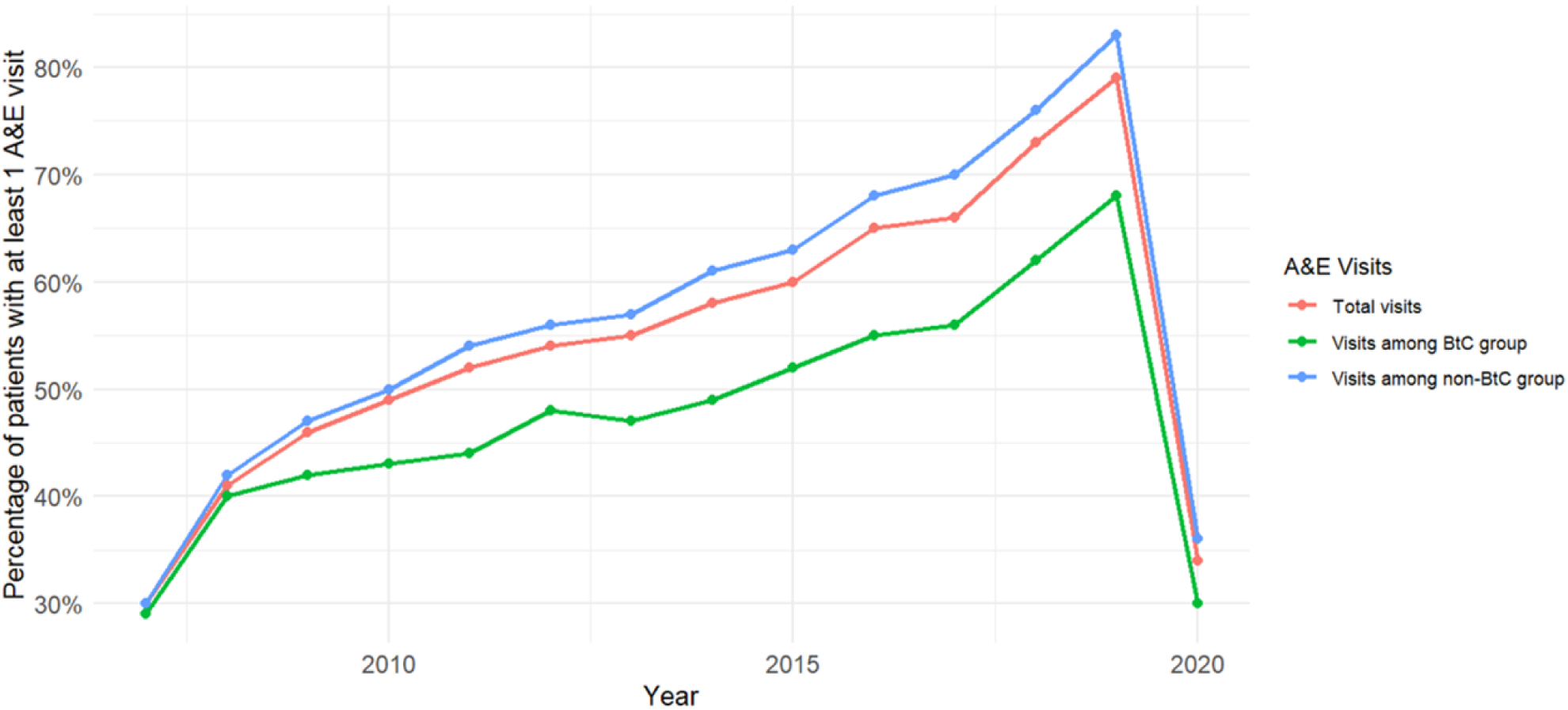
Time trends of A&E attendance by BtC status

#### 4. All-cause and cause-specific outpatient attendance and inpatient admissions

Figure 6 displays the percentage of patients with at least one inpatient admission per year from 2003 to 2022, stratified by BtC status and admission type. The most notable trend is the increase in inpatient admissions for physical health reasons from 2015 onwards, particularly among individuals without BtC, reaching over 80% by 2022. Those with BtC also show a rise in admissions for physical ill-health although at a lower level.

**Figure 6:**
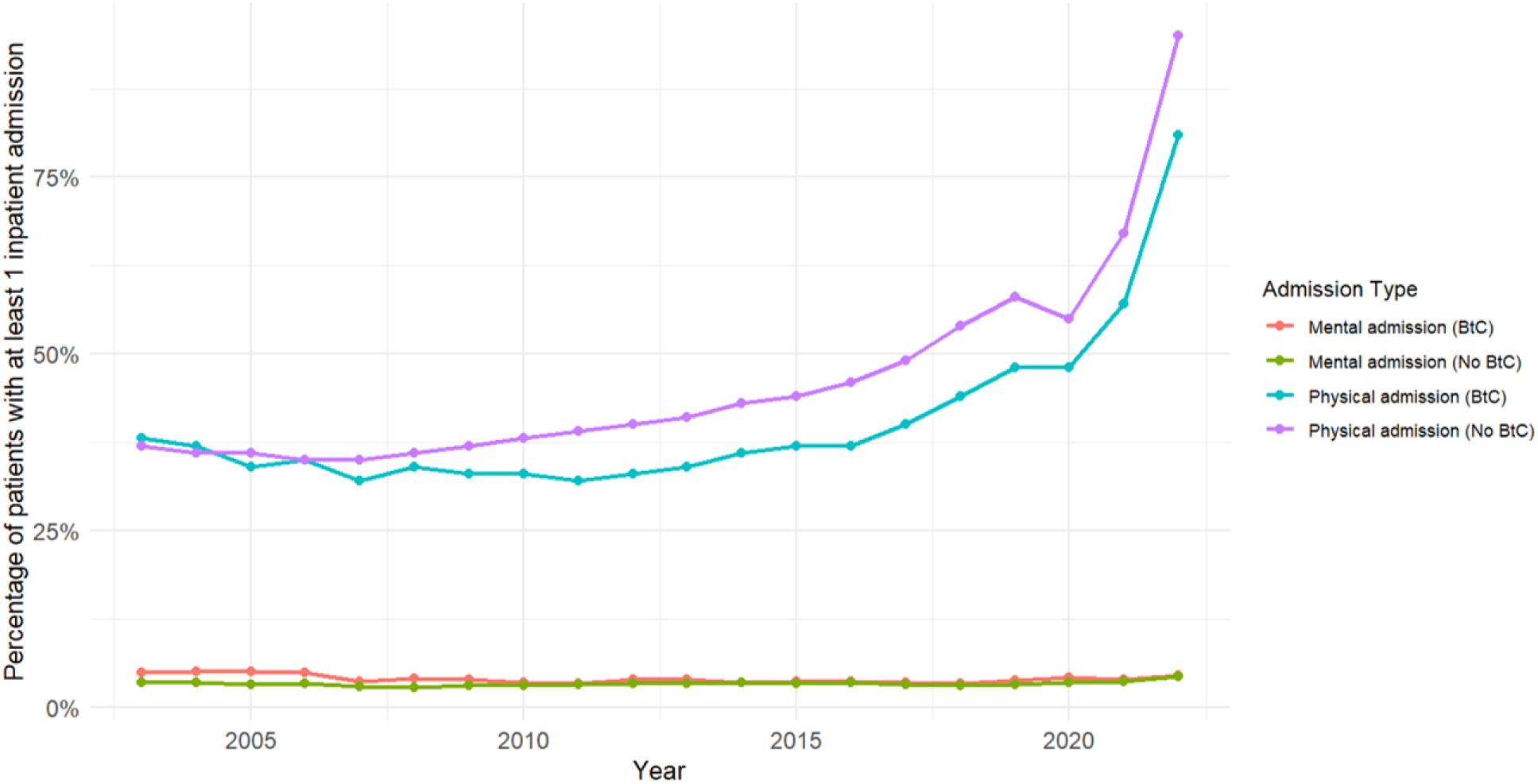
Time trends of inpatient admissions in the cohort by BtC status

In contrast, inpatient admissions for mental health reasons remained relatively low and stable throughout the same period. Both BtC and non-BtC groups showed similar admission rates under 10% throughout the study period.

Figure 7 displays the percentage of patients with at least one outpatient attendance per year from 2003 to 2022, stratified by BtC status and admission type. Physical health outpatient attendance among those with BtC rose from 30% at cohort inception to 49% by 2019. There was a slight decline after 2019, possibly reflecting the impact of the COVID-19 pandemic on in-person consultations. Physical outpatient attendance among those without BtC also increased but the rates were consistently lower than the BtC group.

**Figure 7:**
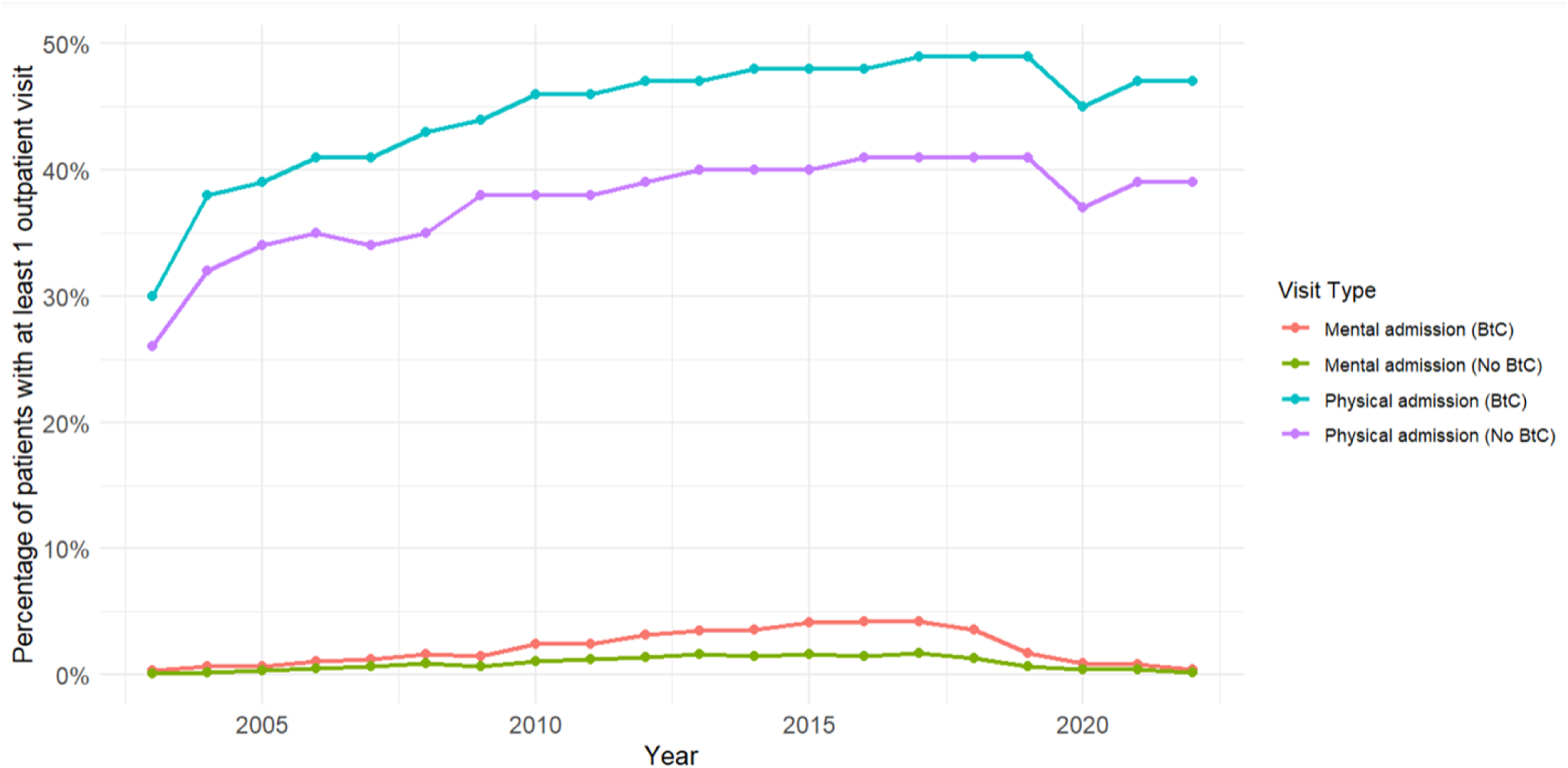
Time trends of outpatient attendance in the cohort by BtC status

Mental health outpatient attendance rates among those with BtC gradually increased from 0.3% at cohort inception to 4.2% by 2017. Mental health outpatient attendance among those without BtC was below 2% throughout the study period.

### Associations of BtC and physical and mental health service use

Table 3 shows that adults with BtC have significantly higher odds of using mental health outpatient services (Model 2: OR: 1.81; 95% CI: 1.71 to 1.93). The effect is reduced after adjusting for mental and physical health comorbidities but remains statistically significant (Model 4: OR: 1.42; 95% CI: 1.33 to 1.52).

**Table 3:**
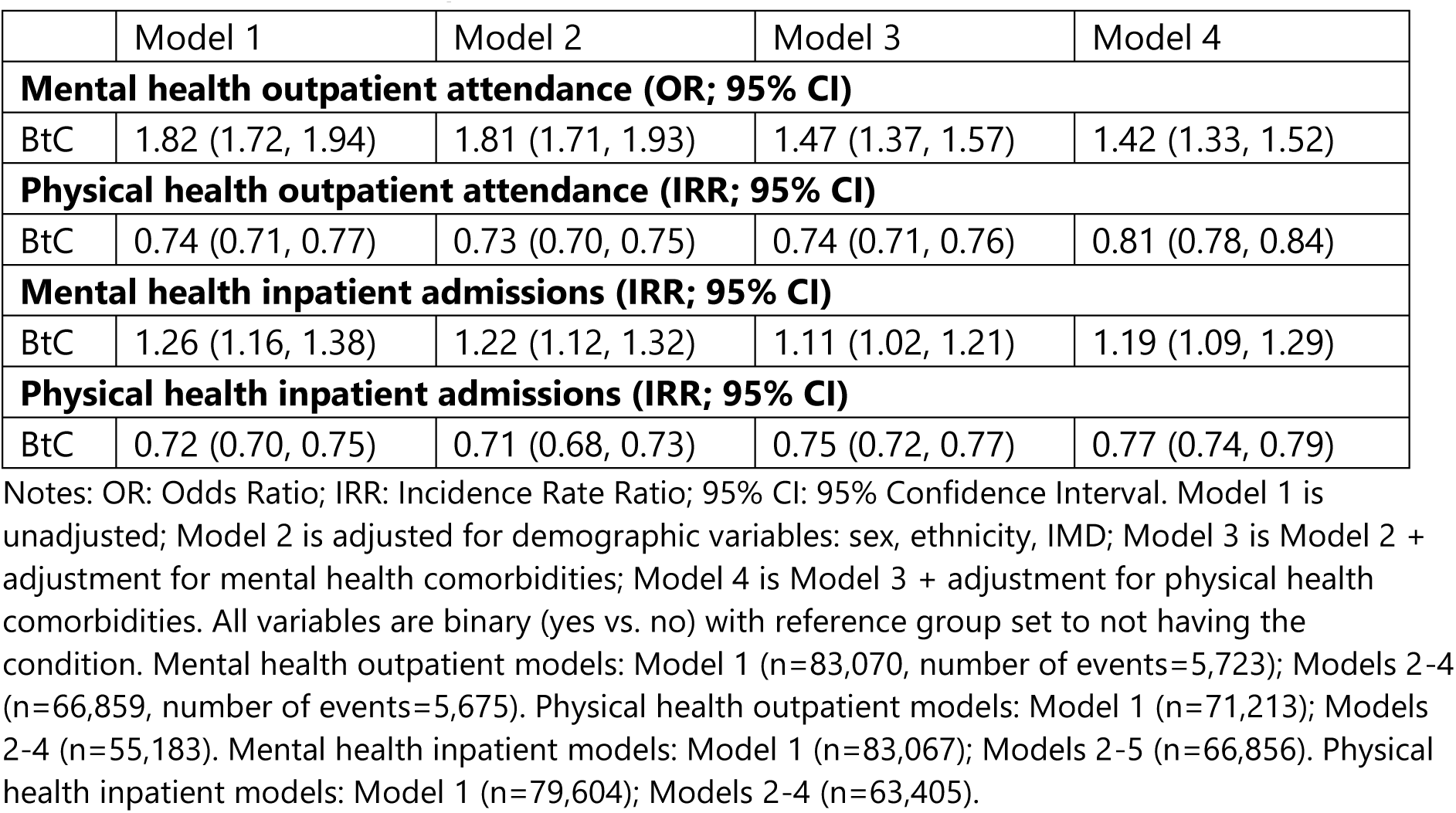
Models assessing the associations between BtC and health service use

BtC is associated with lower rates of physical health outpatient attendance (Model 4: IRR: 0.81; 95% CI: 0.78 to 0.84) but higher rates of mental health inpatient admissions (Model 4: IRR: 1.19; 95% CI: 1.09 to 1.29), even after full adjustment.

Adults who display BtC have consistently lower rates of physical health inpatient admissions than the non-BtC group. The association remains statistically significant after adjusting for demographic characteristics and comorbidities (Model 4: IRR: 0.77; 95% CI: 0.74 to 0.79).

Full estimates for mental and physical health comorbidities are shown in Supplementary Table 1.

#### 5. All-cause mortality and BtC associations

Looking at Table 4, Models 1 and 2 show a small but statistically significant reduction in mortality for those with BtC compared to those without BtC (HR: 0.95; 95% CI: 0.92 to 0.98). This is supported by the Kaplan-Meier curve (Figure 8). Models 3-4, after adjusting for mental and physical health comorbidities, show a weaker and non-significant association, suggesting that health comorbidities explain much of the difference. Despite the high proportions of bipolar disorder, psychoses and schizophrenia observed in Table 1, SMI is less likely to influence mortality (Model 4, HR: 0.93; 95% CI: 0.90 to 0.96). In contrast, neurodevelopmental conditions, anxiety, depression and epilepsy are more likely to influence mortality (Table 4).

**Table 4:**
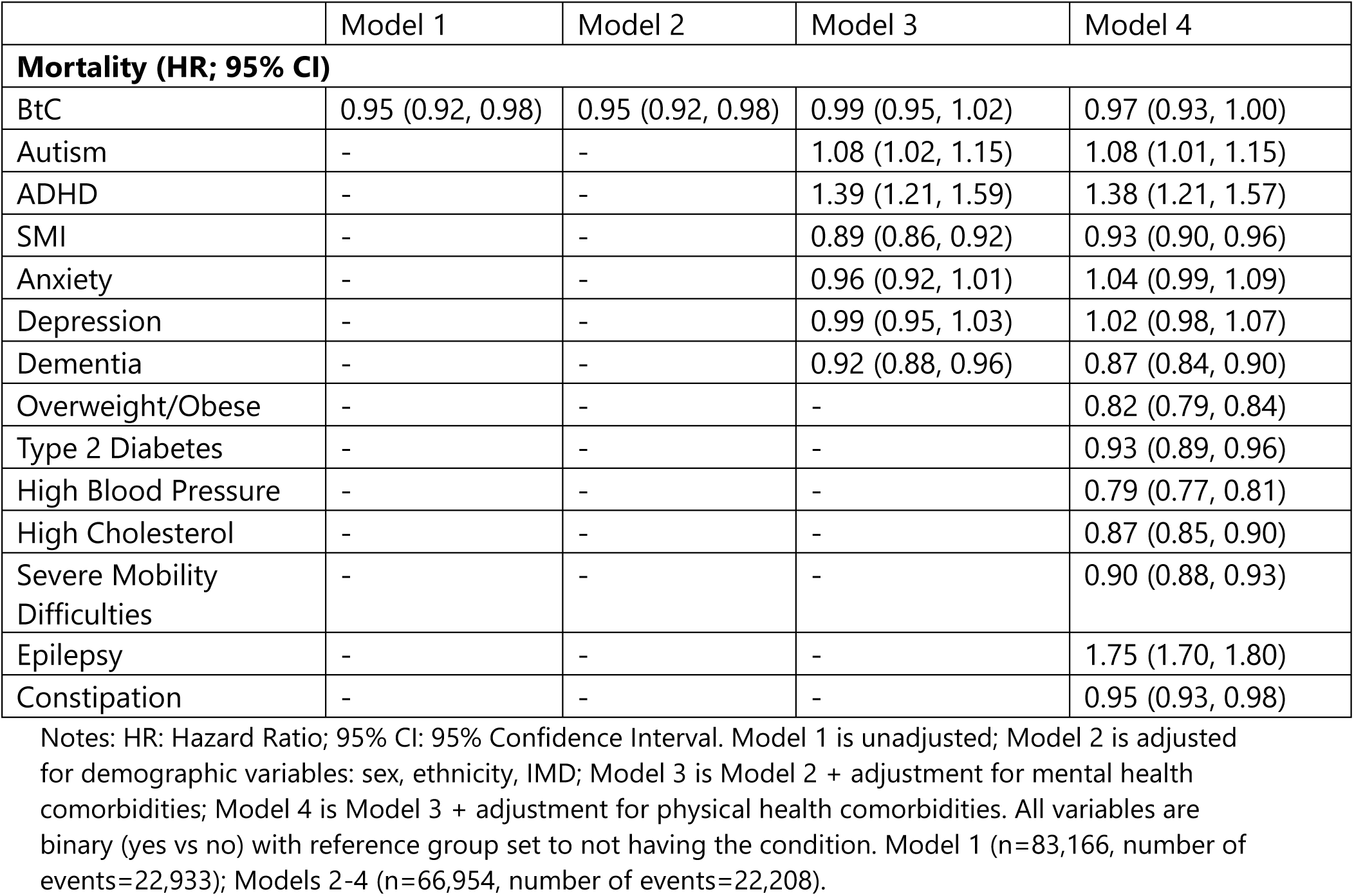
Models assessing the association between BtC and mortality

|  | Model 1 | Model 2 | Model 3 | Model 4 |
| --- | --- | --- | --- | --- |
| **Mortality (HR; 95% CI)** |  |  |  |  |
| BtC | 0.95 (0.92, 0.98) | 0.95 (0.92, 0.98) | 0.99 (0.95, 1.02) | 0.97 (0.93, 1.00) |
| Autism | - | - | 1.08 (1.02, 1.15) | 1.08 (1.01, 1.15) |
| ADHD | - | - | 1.39 (1.21, 1.59) | 1.38 (1.21, 1.57) |
| SMI | - | - | 0.89 (0.86, 0.92) | 0.93 (0.90, 0.96) |
| Anxiety | - | - | 0.96 (0.92, 1.01) | 1.04 (0.99, 1.09) |
| Depression | - | - | 0.99 (0.95, 1.03) | 1.02 (0.98, 1.07) |
| Dementia | - | - | 0.92 (0.88, 0.96) | 0.87 (0.84, 0.90) |
| Overweight/Obese | - | - | - | 0.82 (0.79, 0.84) |
| Type 2 Diabetes | - | - | - | 0.93 (0.89, 0.96) |
| High Blood Pressure | - | - | - | 0.79 (0.77, 0.81) |
| High Cholesterol | - | - | - | 0.87 (0.85, 0.90) |
| Severe Mobility Difficulties | - | - | - | 0.90 (0.88, 0.93) |
| Epilepsy | - | - | - | 1.75 (1.70, 1.80) |
| Constipation | - | - | - | 0.95 (0.93, 0.98) |

Notes: HR: Hazard Ratio; 95% CI: 95% Confidence Interval. Model 1 is unadjusted; Model 2 is adjusted for demographic variables: sex, ethnicity, IMD; Model 3 is Model 2 + adjustment for mental health comorbidities; Model 4 is Model 3 + adjustment for physical health comorbidities. All variables are binary (yes vs no) with reference group set to not having the condition. Model 1 (n=83,166, number of events=22,933); Models 2-4 (n=66,954, number of events=22,208).

**Figure 8:**
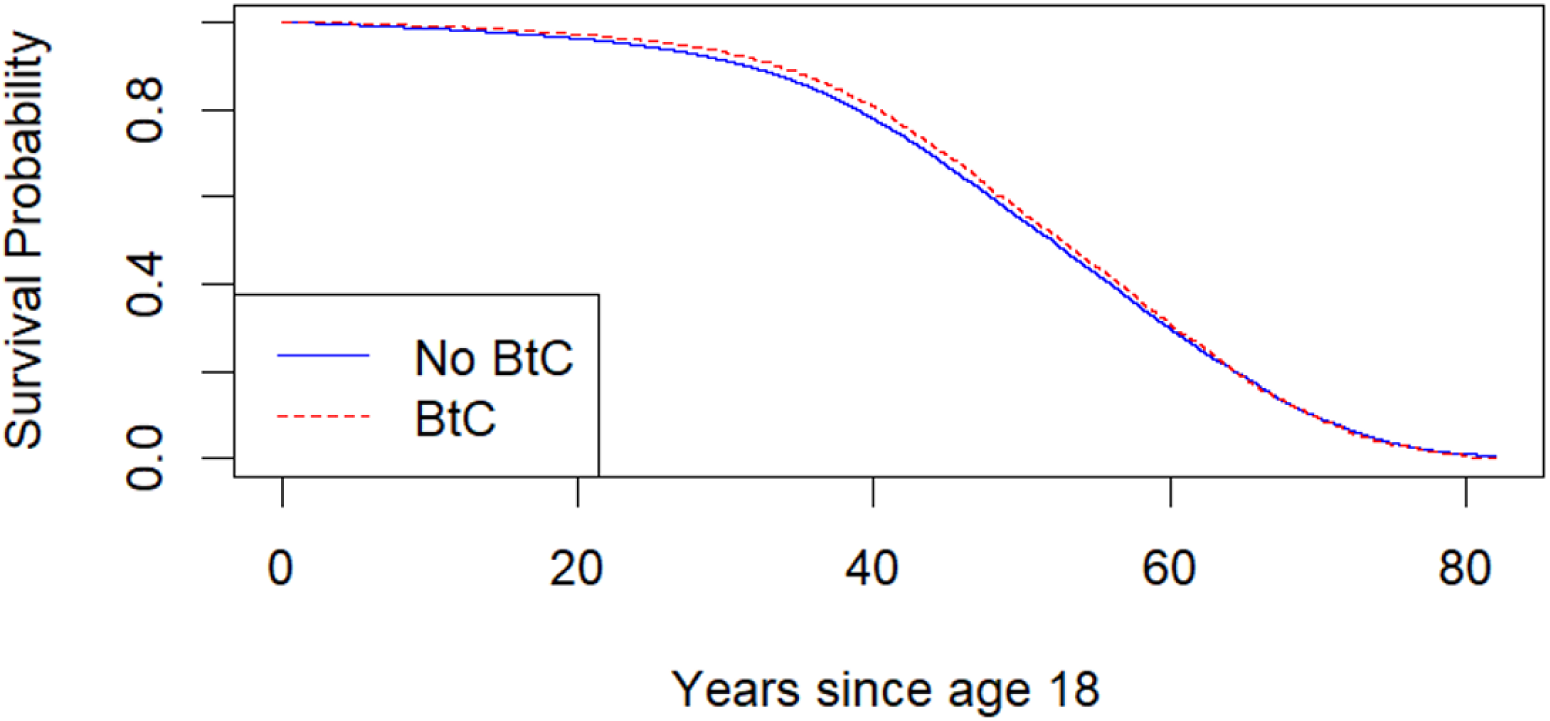
Kaplan-Meier Curve by BtC status (unadjusted)

## DISCUSSION

### Main findings

Adults with intellectual disabilities who display BtC had markedly higher rates of both physical and mental health comorbidities and neurodevelopmental disorders. They also had more recorded psychotropic medications and non-pharmacological interventions compared to those without BtC. 40.5% of those prescribed antipsychotics had no recorded severe mental illness or BtC. BtC was not independently associated with increased mortality, but comorbid neurodevelopmental disorders, common mental disorders, and epilepsy raised mortality risk. As expected, BtC predicted increased use of both inpatient and outpatient mental health services but, counterintuitively, not physical health services.

### Strengths and limitations of the study

This is the first study, to our knowledge, to explore trajectories of health outcomes in a high-need population using a representative, population-based sample with linked secondary care data spanning two decades—allowing for robust longitudinal analysis. Use of clinician-verified code lists for BtC and comorbidities, alongside a wide range of covariates, strengthened the study’s validity. BtC have major implications for individuals with intellectual disabilities and their families and therefore, investigating long term prognosis can support personalised care, improve symptom recording and management, and inform service planning.

Several limitations should be noted. Firstly, clinical coding in electronic health records may not fully capture BtC or mental illness diagnoses, introducing potential misclassification. Secondly, the significant finding of a reduction in mortality among those with BtC in the unadjusted models may be influenced by immortal time bias [19], as individuals could only receive a BtC code if they survived long enough to obtain it, thus gaining additional survival time. Thirdly, non-pharmacological interventions were likely under-recorded, as these are often not shared with GPs and research indicates that psychoeducation is the most frequently offered support in mental health services among those with intellectual disabilities, with 36% offering no named intervention in the past year [20]. Additionally, not all practices had HES linkage, limiting the secondary care analysis sample. Also, we could not stratify by level of intellectual disabilities due to inconsistent recording. Lastly, the observational design limits causal inference, with residual confounding possible.

### Findings in context

The lower contact of physical health service use among individuals with BtC is concerning, especially given the substantial rates of psychotropics prescribing and the relatively higher offer of AHCs [21]. Persistent barriers—such as difficulty accommodating individuals with BtC in physical health settings, diagnostic overshadowing [22], or the episodic nature of BtC—may contribute to this disconnect between need and service access.

Our results reflect a high burden of mental ill-health and neurodevelopmental comorbidities among adults with intellectual disabilities—particularly those with BtC—consistent with prior research [23]. International studies [24, 25] similarly show sustained psychiatric morbidity in this group. While antipsychotic prescribing without a recorded SMI has declined compared to earlier reports [7, 26], it remains prevalent, possibly reflecting improved SMI recognition or changing prescribing thresholds.

Contrary to earlier CPRD studies showing elevated mortality linked to intellectual disabilities [27], we did not find BtC to be independently associated with higher mortality. The earlier cohort (2010–2014) predated key care improvement initiatives—such as the STOMP campaign (Stopping the Overmedication of People with intellectual disabilities) [28]—which may have contributed to more recent health gains.

### Possible explanations and implications for clinicians and policymakers

The growing prevalence of clinical comorbidities in the cohort over time is creating a chronic disease burden for people with intellectual disabilities. Importantly, BtC only drives mental health outpatient attendance and hospitalisations but not physical health input despite national standards recommending physical comorbidity investigation as an early step in the diagnostic pathway [21]. This discrepancy suggests a misalignment between policy and practice and calls for further tailoring of approaches for those with complex behavioural and health needs to improve personalised care for a costly and common mental health condition.

Applying the International Classification of Functioning, Disability and Health (ICF) framework [29] helps conceptualise BtC not merely as a symptom of mental illness but as the outcome of interactions between impairments, environmental barriers, and personal factors. Adults with BtC often face chronic illness and multimorbidity, which heightens vulnerability and complicates care. The high rate of mental health service use may reflect a focus on impairment-based care, while the lack of physical health engagement suggests that other domains of functioning (e.g., participation and access) are not being consistently addressed. The ICF model supports a more holistic and context-sensitive approach, promoting integrated care that combines physical health monitoring, mental health support and non-pharmacological strategies. Such approaches are crucial to achieving equitable health outcomes and enhancing quality of life for people with BtC.

### Unanswered questions and future research

Recent policy changes—such as the removal of AHC targets from the Quality and Outcomes Framework (QOF) [30]—risk de-prioritising these checks, especially in overstretched practices. Additionally, the absence of references to hard-to-reach populations in the latest NHS 10-Year Plan [31] raises concerns that hard-won progress in the care of people with intellectual disabilities including those who display BtC may be reversed. Enhanced data on BtC severity, frequency, context and interventions received promoted by the specialist community services would aid in stratifying risk and guiding preventative strategies. Future research should monitor how policy shifts affect access, treatment delivery, and health equity. This is essential to determine whether reforms are translating into meaningful improvements in outcomes for this high-risk and underserved population.

## Data Availability

CPRD data are not publicly available. However, any other data related to the study (e.g., code lists) can be made available upon reasonable request.

## Author Contributions

Conceptualisation (SD)

Data curation (ALL)

Formal analysis (MJ, LM, AS)

Funding acquisition (AH)

Methodology (ALL)

Project administration (AH, MJ)

Resources (AH)

Software (DR, AS)

Supervision (AH, LM)

Writing original draft (MJ, AH, LM)

Writing review and editing (ALL)

## Transparency Declaration

I, Dr Memta Jagtiani, affirm that the manuscript is an honest, accurate, and transparent account of the study being reported; that no important aspects of the study have been omitted; and that any discrepancies from the study as originally planned (and, if relevant, registered) have been explained.

