## Supplementary Table 1 for "Persistent health inequalities over 20 years among adults with intellectual disabilities who display behaviours that challenge: Evidence from English primary care records"

**Supplementary Table 1: Full estimates for the models assessing the associations between BtC and physical and mental health service use**

| | Model 1 | Model 2 | Model 3 | Model 4 |
|---|---|---|---|---|
| **Mental health outpatient attendance (OR; 95% CI)** | | | | |
| BtC | 1.82 (1.72, 1.94) | 1.81 (1.71, 1.93) | 1.47 (1.37, 1.57) | 1.42 (1.33, 1.52) |
| Autism | | | 1.55 (1.44, 1.66) | 1.57 (1.46, 1.69) |
| ADHD | | | 1.45 (1.31, 1.62) | 1.49 (1.34, 1.66) |
| SMI | | | 1.86 (1.74, 1.98) | 1.83 (1.72, 1.96) |
| Anxiety | | | 1.02 (0.93, 1.12) | 1.02 (0.93, 1.12) |
| Depression | | | 0.97 (0.90, 1.06) | 0.97 (0.90, 1.06) |
| Dementia | | | 1.46 (1.30, 1.63) | 1.41 (1.25, 1.58) |
| Overweight/Obese | | | | 1.07 (1.00, 1.13) |
| Type 2 Diabetes | | | | 0.98 (0.92, 1.05) |
| Overweight, Obese or Diabetic | | | | - |
| High Blood Pressure | | | | 1.08 (0.99, 1.19) |
| High Cholesterol | | | | 1.14 (1.07, 1.22) |
| Severe Mobility Difficulties | | | | 1.00 (0.91, 1.09) |
| Epilepsy | | | | 1.18 (1.10, 1.26) |
| Constipation | | | | 0.95 (0.89, 1.02) |
| **Physical health outpatient attendance (IRR; 95% CI)** | | | | |
| BtC | 0.74 (0.71, 0.77) | 0.73 (0.70, 0.75) | 0.74 (0.71, 0.76) | 0.81 (0.78, 0.84) |
| Autism | | | 1.45 (1.40, 1.50) | 1.36 (1.31, 1.41) |
| ADHD | | | 1.63 (1.53, 1.73) | 1.51 (1.42, 1.60) |
| SMI | | | 1.10 (1.07, 1.14) | 1.20 (1.16, 1.23) |
| Anxiety | | | 0.81 (0.78, 0.85) | 0.90 (0.87, 0.94) |
| Depression | | | 0.91 (0.87, 0.95) | 0.92 (0.88, 0.95) |
| Dementia | | | 0.73 (0.69, 0.76) | 0.81 (0.76, 0.85) |
| Overweight/Obese | | | | 0.89 (0.87, 0.92) |
| Type 2 Diabetes | | | | 0.73 (0.71, 0.76) |
| Overweight, Obese or Diabetic | | | | - |
| High Blood Pressure | | | | 1.48 (1.42, 1.55) |
| High Cholesterol | | | | 0.61 (0.59, 0.62) |
| Severe Mobility Difficulties | | | | 0.96 (0.92, 1.00) |
| Epilepsy | | | | 1.00 (0.97, 1.03) |
| Constipation | | | | 0.85 (0.83, 0.88) |
| **Mental health inpatient admissions (IRR; 95% CI)** | | | | |
| BtC | 1.26 (1.16, 1.38) | 1.22 (1.12, 1.32) | 1.11 (1.02, 1.21) | 1.19 (1.09, 1.29) |
| Autism | | | 1.56 (1.42, 1.72) | 1.52 (1.38, 1.67) |
| ADHD | | | 1.86 (1.61, 2.17) | 1.76 (1.52, 2.05) |
| SMI | | | 2.59 (2.40, 2.80) | 3.10 (2.86, 3.35) |

| | | | | |
|---|---|---|---|---|
| Anxiety | | | 0.79 (0.70, 0.88) | 0.87 (0.78, 0.97) |
| Depression | | | 0.81 (0.73, 0.89) | 0.85 (0.78, 0.94) |
| Dementia | | | 0.42 (0.36, 0.50) | 0.49 (0.42, 0.58) |
| Overweight/Obese | | | | 0.71 (0.65, 0.76) |
| Type 2 Diabetes | | | | 0.75 (0.69, 0.81) |
| Overweight, Obese or Diabetic | | | | - |
| High Blood Pressure | | | | 1.41 (1.26, 1.59) |
| High Cholesterol | | | | 0.55 (0.51, 0.60) |
| Severe Mobility Difficulties | | | | 0.57 (0.51, 0.64) |
| Epilepsy | | | | 1.34 (1.24, 1.46) |
| Constipation | | | | 0.95 (0.87, 1.03) |
| **Physical health inpatient admissions (IRR; 95% CI)** | | | | |
| BtC | 0.72 (0.70, 0.75) | 0.71 (0.68, 0.73) | 0.75 (0.72, 0.77) | 0.77 (0.74, 0.79) |
| Autism | | | 0.86 (0.82, 0.89) | 0.84 (0.81, 0.87) |
| ADHD | | | 1.12 (1.05, 1.18) | 1.10 (1.04, 1.17) |
| SMI | | | 1.03 (1.00, 1.06) | 1.11 (1.07, 1.14) |
| Anxiety | | | 0.80 (0.77, 0.84) | 0.88 (0.84, 0.92) |
| Depression | | | 0.93 (0.89, 0.96) | 0.93 (0.90, 0.97) |
| Dementia | | | 0.75 (0.71, 0.80) | 0.76 (0.72, 0.81) |
| Overweight/Obese | | | | 0.73 (0.71, 0.75) |
| Type 2 Diabetes | | | | 0.90 (0.87, 0.93) |
| Overweight, Obese or Diabetic | | | | - |
| High Blood Pressure | | | | 1.71 (1.64, 1.79) |
| High Cholesterol | | | | 0.60 (0.58, 0.62) |
| Severe Mobility Difficulties | | | | 1.13 (1.09, 1.17) |
| Epilepsy | | | | 1.18 (1.14, 1.21) |
| Constipation | | | | 0.97 (0.94, 1.00) |

Notes: OR: Odds Ratio; IRR: Incidence Rate Ratio; 95% CI: 95% Confidence Interval. Model 1 is unadjusted; Model 2 is adjusted for demographic variables: sex, ethnicity, IMD; Model 3 is Model 2 + adjustment for mental health comorbidities; Model 4 is Model 3 + adjustment for physical health comorbidities. All variables are binary (yes vs. no) with reference group set to not having the condition. Mental health outpatient models: Model 1 (n=83,070, number of events=5,723); Models 2-4 (n=66,859, number of events=5,675). Physical health outpatient models: Model 1 (n=71,213); Models 2-4 (n=55,183). Mental health inpatient models: Model 1 (n=83,067); Models 2-5 (n=66,856). Physical health inpatient models: Model 1 (n=79,604); Models 2-4 (n=63,405).
